# Relational One Health: a more-than-biomedical framework for more-than-human health, and lessons learned from Brazil, Ethiopia, and Israel

**DOI:** 10.1101/2023.10.10.23296827

**Authors:** Julianne Meisner, Hilary McLeland-Wieser, Elizabeth E. Traylor, Barak Hermesh, Tabata Berg, Amira Roess, Lauren Van Patter, Anat Rosenthal, Nadav Davidovitch, Peter M. Rabinowitz

## Abstract

The One Health conceptual framework envisions human, animal, and environmental health as interconnected. This framework has achieved remarkable progress in the control of zoonotic diseases, but it commonly neglects the environmental domain, implicitly prioritizes human life over the life of other beings, and fails to consider the political, cultural, social, historical, and economic contexts that shape the health of multispecies collectives. We have developed a novel theoretical framework, Relational One Health, which expands the boundaries of One Health, clearly defines the environmental domain, and provides an avenue for engagement with critical theory. We present a systematic literature review of One Health frameworks to demonstrate the novelty of Relational One Health, and to orient it with respect to other critically-engaged frameworks for One Health. Our results indicate that while Relational One Health complements several earlier frameworks, these other frameworks are either not intended for research, or for narrow sets of research questions. We then demonstrate the utility of Relational One Health for One Health research through case studies in Brazil, Israel, and Ethiopia. Empirical research which is grounded in theory can speak collectively, increasing the impact of individual studies and the field as a whole. One Health is uniquely poised to address several wicked challenges facing the 21^st^ century—climate change, pandemics, neglected zoonoses, and biodiversity collapse—and a unifying theoretical tradition is key to generating the evidence needed to meet these challenges.

**HIGHLIGHTS:**

- One Health views human, animal, and environmental health as interconnected
- Biomedical reductionism in One Health has resulted in a focus on human health threats from animals
- The environmental domain and more-than-biomedical contexts are commonly ignored in One Health
- Relational One Health is a new theoretical framework which addresses these limitations
- This theoretical framework is relevant to all One Health research, increasing the field’s impact

## BACKGROUND

The COVID-19 pandemic has increased interest in institutionalizing a One Health approach to prevent future pandemics. This is rooted in a growing consensus that zoonotic emergence is caused by human-environment relations grounded in colonial-capitalism and resulting in habitat loss and climate change, and thus likely to continue. At the same time, social movements accelerated by the pandemic have collided with the undeniable fact that infectious diseases are not just biomedical phenomena: rather, the transmission and impacts of pathogens follow long-established gradients of social difference. These gradients are politically, historically, and geographically contextual: their deep historical roots have been perpetuated and upheld by social, economic, and political structures, and today manifest as socioeconomic inequities. Within One Health, white papers and reports published by multilateral agencies and commissions have reflected this growing acceptance that One Health cannot occupy a solely biomedical position. Yet to our knowledge, this awareness is not coupled with a suitable theoretical framework for One Health research.

The One Health High Level Expert Panel (OHHLEP) was convened in 2021 by the Quadripartite partners—Food and Agriculture Organisation of the UN (FAO), UN Environment Programme (UNEP), World Health Organisation (WHO) and World Organization for Animal Health (WOAH)—to support development of the Quadripartite’s One Health Joint Plan of Action. OHHLEP’s first task was to develop a new, action-oriented definition of One Health, from the many dozens of definitions in circulation (1). Indeed, OHHLEP’s new definition is certainly more-than-biomedical, supported by key underlying principles which emphasize “sociopolitical and multicultural parity, socioecological equilibrium, and epistemological equity.” However, it is intended to guide policy, not research. Conversely, the Lancet One Health Commission published a new definition of One Health in 2020 focused on research (2). While there are more-than-biomedical dimensions to the Commission’s definition, they do not extend to the knowledge generating process itself, rather beginning when research is translated into policy: “…evidence generation must be used to drive context-driven governance, progressive policy, and legislation that are sensitive to gender, community, equity, and ethics.”

### Box 1: Key definitions

**One Health*:** A collaborative approach to attain optimal health for people, domestic animals, wildlife, plants, and our environment. **EcoHealth*:** A movement which recognizes the inextricable dynamic linkages between the health of all species and their environments. **Planetary Health*:** Achievement of the highest attainable standard of health, wellbeing, and equity worldwide through judicious attention to human systems and the Earth’s natural systems that define the safe environmental limits within which humanity can flourish. **Ecosystems:** Communities of interacting beings, biotic and abiotic. **Environment:** The biophysical, social, cultural, political, historical, and economic contexts (systems, structures, and circumstances) in which humans, animals, and ecosystems exist. *\*Adapted from Lerner and Berg, 2017*

Beyond biomedical reductionism, One Health has also been criticized for failing to define the environmental domain, or ignoring it entirely. The original vision for One Health integrated human, animal, and environmental health with other social science disciplines: a truly holistic “more-than-human” approach. In practice, One Health has largely stopped at integrating human and animal health, focused on zoonotic diseases within the veterinary and healthcare sectors. Further, donor priorities have led to an implicit hierarchy which places humans over other beings. Animals are often viewed as “exposures” or threats to human health, rather than health bearers in their own regard, and exceptions to this are usually framed in terms of agricultural productivity and economic losses, and thus still reflect human priorities. It is quietly accepted that this neglect of the environmental domain in One Health motivated the advent of the Planetary Health movement in 2014 (3), however Planetary Health takes an anthropocentric view of health, and thus aligns more with Global Health than with One Health. EcoHealth, by contrast, is interested in the wellbeing of all living creatures. There is significant overlap between EcoHealth and One Health, however One Health focuses on the health of individuals, while EcoHealth focuses on aggregations (4) (Box 1). In 2017 the COHERE guidelines were published to encourage better representation of the environmental domain in One Health papers (5), and in 2020 the Tripartite (WHO, FAO WOAH) expanded to include the UNEP as the new Quadripartite. These shifts are accompanied by attempts to better define the environment (plants, soil, water, etc.) within One Health policy statements and white papers. We argue, however, that One Health’s conceptualization of the environment has yet to crystallize, despite these efforts. The environment can be defined very broadly—all elements of the physical, cultural, social, and political milieu—or narrowly—the immediate built environment and its hazards—and thus this omission is non-trivial.

We introduce Relational One Health (Figure 1) as a novel, critically-engaged theoretical framework which seeks to clarify how the environment is conceptualized within One Health, challenge One Health researchers to think beyond biomedical dimensions and determinants of multispecies health, and subvert the implicit prioritization of humans over other living beings. Under this framework, the distribution of health is a collective over and within humans, non-human animals (“animals”, for simplicity), and ecosystems, each of which are health bearers. Ecosystems subsume animals, and animals subsume humans, reflecting the relationality between them. These health bearers share a common environment which determines the distribution of health and has social, cultural, historical, political, economic, and biophysical dimensions. This framework revisits One Health’s initial holistic vision, challenging the constraints that have settled around One Health over the past few decades. It also provides foundation for critically-engaged scholarship within One Health, as the environmental domain cannot be fully examined or understood if systems of power and oppression, which ultimately shape the circumstances that determine health, are ignored. We first present our findings from a systematic review of One Health frameworks. We then detail the Relational One Health theoretical framework’s assumptions and origins, and conclude by presenting brief case studies from Brazil, Israel, and Ethiopia, from the lens of the Relational One Health theoretical framework.

**Figure 1:**
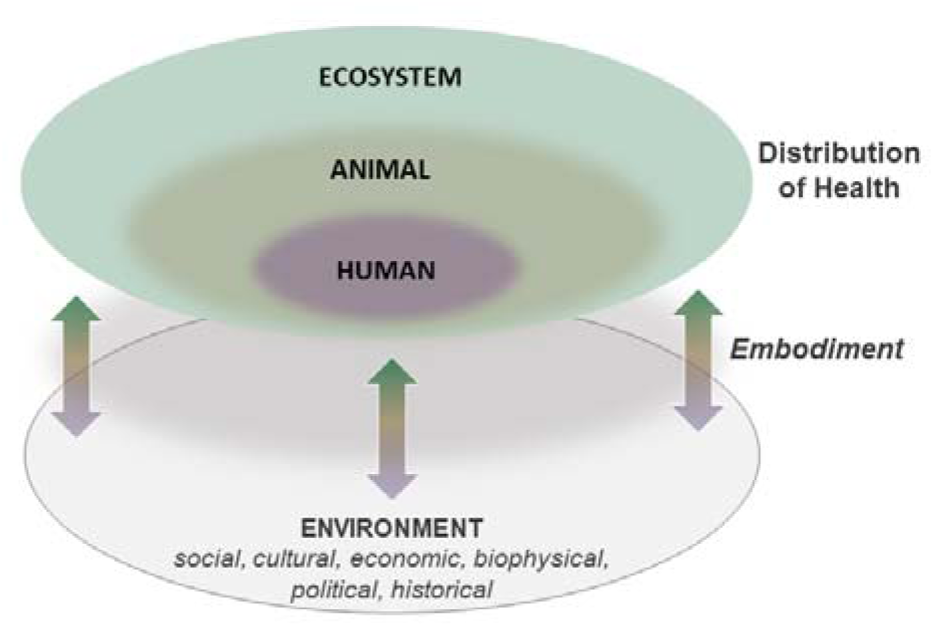
The Relational One Health theoretical framework. The distribution of health is a collective over and within human, animal, and ecosystem health, with each component recognized as health bearers. Humans are subsumed by animals, and animals are subsumed by ecosystems. These biotic and abiotic beings share a common environment which has economic, biophysical, political, historical, and social, cultural dimensions. This shared environment influences health through the active, reciprocal process of embodiment, through which health bearers interface with and influence their environment.

## METHODS

This PRISMA checklist was used to guide this review and its presentation (6).

### Objective

The objective of this systematic review was to evaluate the need for a novel, politically-engaged theoretical framework for One Health, effectively ensuring that Relational One Health doesn’t duplicate other efforts. We sought to find One Health conceptual models or theoretical frameworks, hereafter referred to as “frameworks,” and evaluate the extent to which they engage with more-than-biomedical definitions of health.

### Search strategy

Searches were conducted in PubMed and Web of Science. Results were imported into Research Rabbit (7) to identify similar publications not found in these databases. Search terms, developed by EJT and JM in collaboration with a health sciences librarian, were:

- PubMed: “one health”[Title/Abstract] AND (“framework”[Text Word] OR “model”[Text Word] OR “theor*”[Text Word])
- Web of Science: ALL=(”One Health” AND (“framework” or “model” or “theor*”))

### Selection of publications

Two authors with expertise in One Health [EET and HMW] selected publications, with a third author [JM] consulted when there was uncertainty whether a publication should be included or excluded. Publication titles were subjected to two rounds of reviews by EJT, with selected publications imported into Research Rabbit between these two rounds, and publications found in Research Rabbit incorporated into the second round of title review. Full-text review was then conducted by HMW and JM to select the final publications to be reviewed according to the following criteria:

Inclusion criteria:

- English language
- Full text available
- Title contains the following terms: (“framework” or “model” or “system”) and (“One Health” or “EcoHealth”)
- Framework reconceptualizes and/or re-theorizes One Health (*what is One Health*)
- Framework includes more-than-biophysical contexts for health

Exclusion criteria:

- Title includes the term “impact”
- Publication focuses on a specific pathogen or disease
- Framework focuses on implementation (*how to do One Health*)

All publications were stored in a shared Zotero folder, with subfolders used to identify database(s) of origin and review fate (inclusion/exclusion).

### Data extraction

Once the final selection of publications had been established, HMW extracted data from each included publication into an Excel matrix with the following fields, meeting bi-weekly with JM to discuss the evolution of her findings and resolve conflicts:

- Metadata: title, first author, year published, journal
- Summary of the framework presented
- Framework is presented as novel conceptualization of One Health [Y/N]
- Framework includes sociocultural contexts for health [Y/N]
- Framework includes political contexts for health [Y/N]
- Framework includes economic contexts for health [Y/N]
- Framework includes other more-than-biophysical contexts for health [free text]
- Framework is critically-engaged [Y/N]

We define critically-engaged frameworks as those which discuss or reference critical theory (anti-racist, decolonial, feminist, posthumanist, and others), typically with a lens towards justice and rights.

In this review, we are seeking to capture the same directionality implied by Relational One Health. For example, we are interested in frameworks which examine political economies as causes of multispecies disease distribution, not frameworks for drawing economic arguments in favor of One Health, or frameworks for identifying policies and institutional structures required for successful implementation of One Health.

## RESULTS

### Literature review

PubMed was searched on February 2^nd^, 2023, and Web of Science on February 23^rd^, 2023, producing 1,171 and 1,294 results, respectively. After the first round of title review, 94 publications remained. These were imported into Research Rabbit, which found 30 additional publications. The second round of title review excluded an additional 95 publications, leaving 29 for full-text review. Following full-text review an additional 10 publications were identified from the reference lists, and an additional 31 were excluded. A total of eight publications presented new definitions, theoretical frameworks, or conceptual models for One Health which considered health as a more-than-biomedical phenomenon (i.e., shaped by more-than-biophysical contexts) (Figure 2). All were published since 2009, with the majority published after 2018.

**Figure 2:**
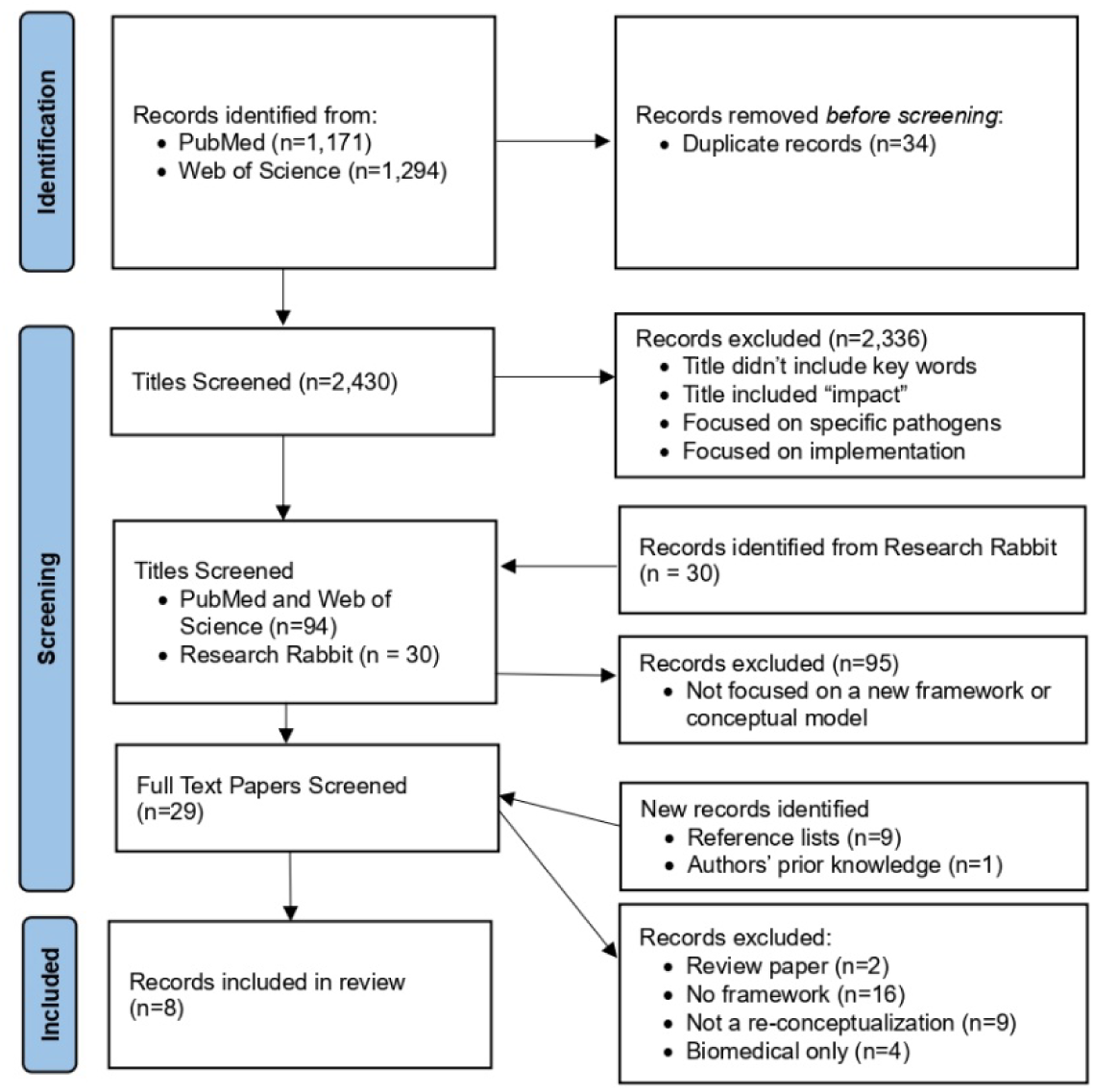
Inclusion flow chart

All publications considered all of the more-than-biomedical contexts we evaluated (sociocultural, political, and economic), thus Table 1 summarizes the framework briefly, and indicates whether the framework was critically-engaged and provided a clear definition of the environmental domain. Of the frameworks labeled as not critically-engaged, several (8,9) contained text which indicated the authors’ openness towards critical theory principles, particularly posthumanism and decolonial theory.

**Table 1:** Summary of publications reviewed.

| Publication | Framework objective and summary | Critically-engaged? | Clear definition of environment? |
| --- | --- | --- | --- |
| Baquero et al. (2021) (9) | Core thesis focuses on marginalizing apparatuses which operate on humans, animals and ecosystems, and identifying structural alternatives for multispecies justice.<br>*<u>Name:</u> One Health of Peripheries* | Yes | Yes; environment is ecosystem health |
| Hardy and Standley (2022) (10) | Identifying commonalities with feminist and Queer theory: decomposing false binaries, challenging reductionism, and examining intersections | Yes | No |
| Lerner and Berg (2014) (11) | Redefining health and questioning the normative connotation behind the term “non-human animals.” | Yes | Yes; environment is ecosystem health |
| Rock et al. (2009) (12) | Uses the idea of syndemics to examine the links between biology and social systems to align with other values for social change and achieve health for all. Focus is on developing prevention and intervention strategies, not examining the determinants of health. | Yes | No |
| Sleeman et al. (2019) (13) | Systems-based harm reduction approach to One Health interventions. | No | Yes; ecosystem is a health bearer, and distinct from the environmental quality |
| Wallace et al (2015) (14) | Empirical framework for studying structural causes of novel pathogen emergence. *<u>Name:</u>* | Yes | Not explicitly; assumed to be |
|  | *Structural One Health* |  | ecosystems |
| Woldehanna and Zimicki (2015) (7) | Framework for examining proximal determinants of human-animal contact (social, cultural, livelihood, etc.). <u>Name:</u> *Expanded One Health* | No | No |
| Zinsstag et al. (2009) (8) | An extension of One Health, EcoHealth, systems biology, and socioecological systems conceptual thinking: <u>Name:</u> *Health in Socio-ecological Systems* | No | Yes; describe a socioecological continuum |

Baquero presents One Health of Peripheries (10), which draws from the perspective of Latin American collective health and more-than-human biopolitics. Central to Baquero’s framework are marginalizing apparatuses, which legitimatize exploitation against living beings on the other side of the margins. Latin American collective health addresses power hierarchies in the social determination of health, but does not consider animals as health bearers and therefore reproduces marginalizing apparatuses that create more-than-human health inequities. Conversely, One Health views animals as health bearers, but largely ignores social processes in both theory and research. Baquero reviews the history of animalization, domestication, colonialization, and slavery to demonstrate that marginalization is a political process which is common not only to humans, but to other animals and the environment, calling for multispecies intersectionality. However, One Health of Peripheries is intended to be a discursive tool, rather than a foundation for research, and no figure or visual is presented.

Hardy & Standley (11) review the symbiotic relationship between One Health and intersectional feminist thought, which originated in Black feminism and has been applied to many overlapping systems of power. General biological classification which was popularized during the Enlightenment era formed the basis for racial classification established during the expansion of Western colonialism. This system of classification led to the historical isolation of health sectors which One Health seeks to address, and to the false binaries that One Health (i.e., animal vs. human) and feminist and Queer theory (i.e., man vs. woman) seek to decompose. This doesn’t mean categories are discarded, but rather their points of intersection are studied and viewed as products of culture, rather than inflexible laws of nature.

The framework presented by Lerner & Berg (12) challenges normative definitions of both health and non-human animals, the latter challenge justified by the huge variety of animal life forms. Health is presented as an individual-level construct: individuals nest in populations, and population-level health is simply the use of statistics to measure the health of individuals that comprise the population (public health for humans, population health/herd health for animals).

Populations then nest in ecosystems, which the authors appear to treat as a health bearer though this is not stated explicitly. A new definition of health that applies to all animals, including humans, is comprised of physiological health, mental health, and balance theory. Similar to Baquero, they consider animals and humans to be part of the same multispecies community. However, Lerner & Berg are focused on definitions, not determinants, of health.

Rock et al. (13) use the concept of a syndemic—two or more afflictions that interact synergistically to result in an excess burden of disease—to re-conceptualize One Health preventions. A syndemic orientation focuses on connections between health problems when developing health policies, aligning with other values for social change to achieve health for all. Using the example of the mental health impacts of widespread animal culls during the 2001 foot and mouth disease outbreak in the UK, the authors demonstrate that prevention strategies should consider not just the need to control infections, but also the economic, cultural, emotional, and political principles that shape the “entwining of biology with social systems.” While this framework is presented as a tool for examining the impacts of policies and interventions and rather than determinants of health, its theoretical foundation couples nicely with the Relational One Health theoretical framework. However, it does not provide an explicit definition of the environment.

Sleeman et al. (14) propose a systems-based harm-reduction approach to One Health interventions, conceptualized through a One Health impact pyramid. At the base of the pyramid are socioeconomic and environmental determinants of health, which act together to underpin decisions and interventions. The authors emphasize that actions should be prioritized here for long-term sustainability, but their examples highlight very proximal risk factors and similarly proximal solutions: e.g., providing alternative sources of income for mining communities during high-risk periods for Marburg transmission so workers don’t enter caves. The environment is clearly defined in this framework: humans, wildlife, and domestic animals are health bearers, which share an environment defined by ecosystem health and environmental quality (presumably air, water, soil, etc.).

Wallace et al.’s Structural One Health (15) critiques analyses of risk factors for pathogen emergence which only examine conditions in geographic proximity to the emergence event, and the implicit assumption of neoliberalism as the status quo in such analyses. In contrast, Structural One Health hypothesizes that global capital accumulation is a key determinant of ecosystem health, and pathogen emergence in turn. This framework empirically formalizes the connections between global circuits of capital, deep-time histories, and cultural infrastructure, which together result in landscape changes and shifts in wildlife, agricultural, and human health. The authors present a schematic for their model in the form of a pyramid, with Structural One Health at the bottom (comprising the broadest context of disease) and emergency medicine at the top, and discuss several applications of the framework which are not yet published. However, Structural One Health is a framework for a particular type of One Health research question (e.g., focused on distal, structural causes), while Relational One Health is intended to be a tool for all One Health investigations (e.g., encompasses structural, distal, and proximal causes), and thus serves a broader purpose.

Woldehanna and Zimicki (8) present a framework for systematically examining the proximate determinants of human-animal contact, with the goal of considering socioecological factors that can contribute to disease emergence at the local level. They refer to this as an “expanded One Health model,” which theorizes that different people living in the same location, affected by the same large-scale drivers, may be at differential risk of spillover due to social factors. Categories of determinants the authors examined include biological characteristics of individuals; social characteristics of individuals, households, and communities, including norms, livelihood systems, and settlement patterns; and at the public policy level, local and international governance and politics.

Finally, Zinsstag et al. (9) present the “Health in Socio-ecological Systems” framework, an extension of One Health, EcoHealth, systems biology, and socioecological systems conceptual thinking. Humans and animals are health bearers existing in a socioecological continuum: animals, including wildlife, are part of the social systems of humans, and part of the environment. The authors present a reciprocal relationship between health and social, cultural, economic, and political contexts: these contexts shape health, and are also determined by health.

### Relational One Health

Despite the promise of One Health, its application has been criticized as reductionist in scope with inter-disciplinary collaborations largely limited to veterinary and healthcare sectors and focused on laboratory methods and surveillance (16). While recent years have seen an increase in more-than-biomedical frameworks for One Health, as reviewed here, these frameworks are either not well-suited to research, or deal only with a specific type of research question. Further, many do not explicitly define the environmental domain (Table 1).

We developed the Relational One Health (Figure 1) theoretical framework to expand the scope of One Health research to include macrosocial determinants of multispecies health, coupled with a clear definition of the environmental domain as distinct from ecosystems. Relational One Health was inspired by several earlier One Health frameworks, particularly Baquero’s One Health of Peripheries (10), which inspired the subsumption in Figure 1, as well as Wallace et al.’s Structural One Health (15) and Hermesh et al.’s Political One Health (17). Relational One Health also draws inspiration from Kreiger’s ecosocial theory of disease distribution (18), which combines social, ecological, and historical context with a postulated mechanism for their influence on health, termed embodiment.

We will now present three case studies which use the Relational One Health theoretical framework to guide research. At the study development stage, this theoretical framework can be used to develop conceptual models, which are then linked to data collection and analysis plans. Later in the research lifecycle, the Relational One Health theoretical framework can be used to contextualize results, triangulate findings with other literature (including outside of the biomedical sciences), and identify directions for future research.

*Case study 1: Emancipatory land rights and arboviruses in Brazil* Emancipatory land rights is defined as secure land control and land access for communities facing dispossession of their land. This is the focus of a research project we are implementing among Quilombola communities in the midwest of Brazil, the descendants of individuals who escaped slavery among Brazil’s regional biomes. Figure 3 presents a conceptual model developed from the Relational One Health theoretical framework to guide this research. Land control is a facet of the environment which itself has political and historical determinants, namely land policy, constitutional rights for Quilombola peoples, and the history of emancipation and rural settlement. Land control influences ecosystem preservation through social and economic intermediates, with weak land rights allowing conflict within and between communities to manifest as ecosystem destruction through capitalist exploitation of the land. This ecosystem destruction increases vectorial capacity and drives arbovirus transmission, an embodiment of land control. Animals, humans, and ecosystems (indicated by colored shading) are all health bearers, and insect vectors are both a component of ecosystems, and also comprise the biophysical environment within which animals and humans exist.

**Figure 3.**
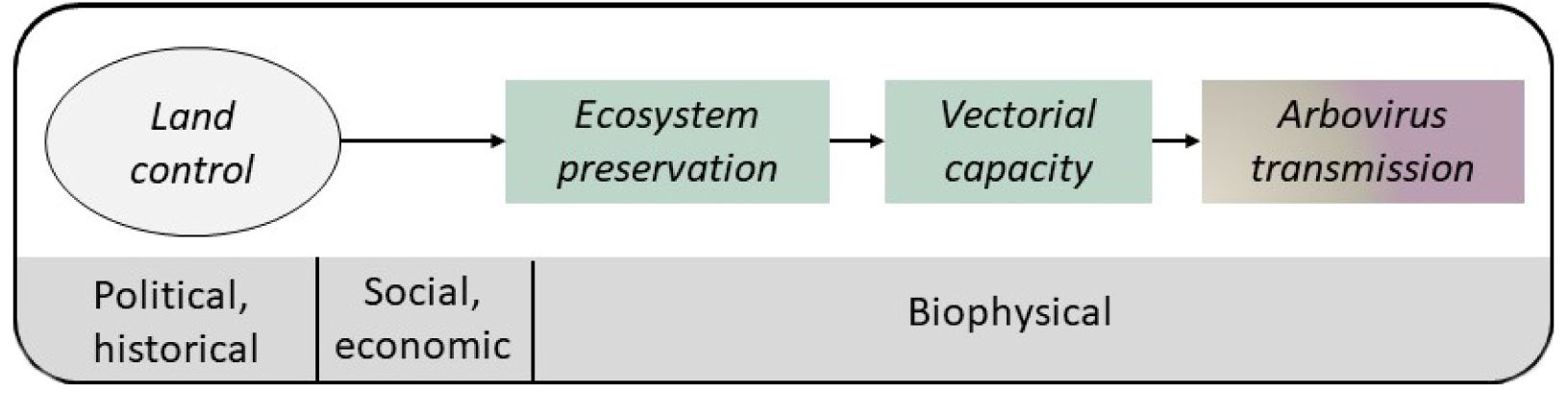
Conceptual model for Case Study 1. Land control is a component of the environment which has political (land policy, Quilombola rights) and historical (emancipation, rural settlement) antecedents, and shapes the health of ecosystems (green shading), and animals and humans (beige/purple gradient), embodied through the transmission of arboviruses. The effect of land control on ecosystem preservation has social (conflicts within and between communities) and economic (capitalist exploitation of land) determinants. The ecosystem (green shading), including the vectors it contains, is both a health bearer and comprises the biophysical environment which shapes animal (beige shading) and human (purple shading) health.

*Case study 2: Distrust and brucellosis in Israel* Case Study 2 examines human-animal contact networks as an intermediary between larger social contexts and zoonotic disease transmission (Figure 4). Specifically, distrust of formal institutions (veterinary, agricultural, public health, economic, etc.) among Bedouin communities in southern Israel is caused by current and historical political, racial/ethnic, and economic contexts. These distal contexts intertwine with more proximal social and economic forces which determine the structure and stability of human-animal and animal-animal contact networks, for instance herd size and husbandry, purchase of animals and animal products from the West Bank, household size, distribution of husbandry tasks within the household, etc. These contact networks in turn facilitate the transmission of pathogens between animals and humans including *Brucella,* a major cause of human and livestock morbidity in Israel.

**Figure 4.**
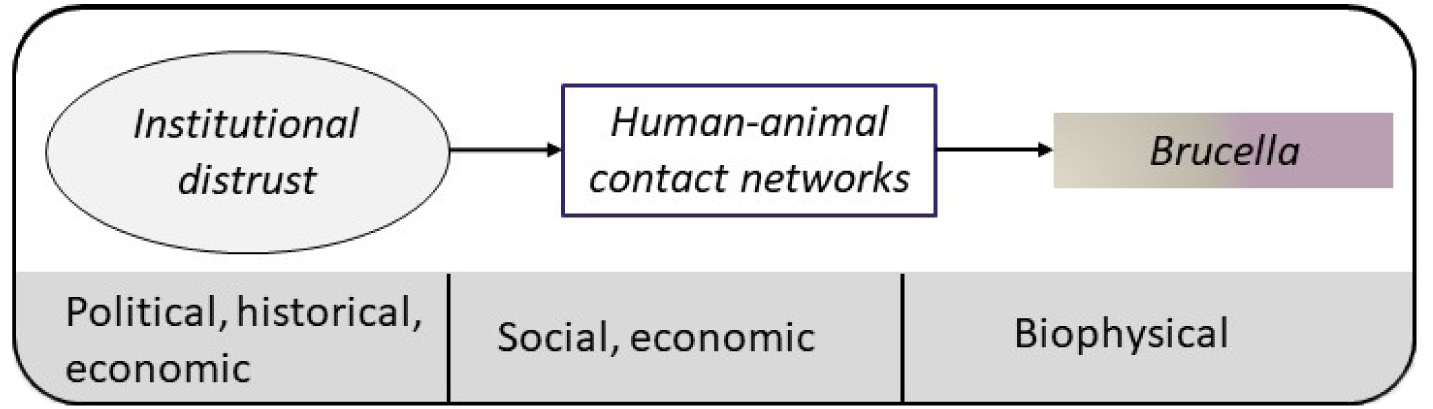
Conceptual model developed for Case Study 2. Among Bedouin communities in southern Israel, political, historical, and economic contexts have resulted in institutional distrust. Institutional distrust, through social and economic intermediates, shapes human-animal contact networks on which pathogens such as *Brucella* spp. are transmitted between humans (purple shading) and food producing animals (beige shading).

**Figure 5.**
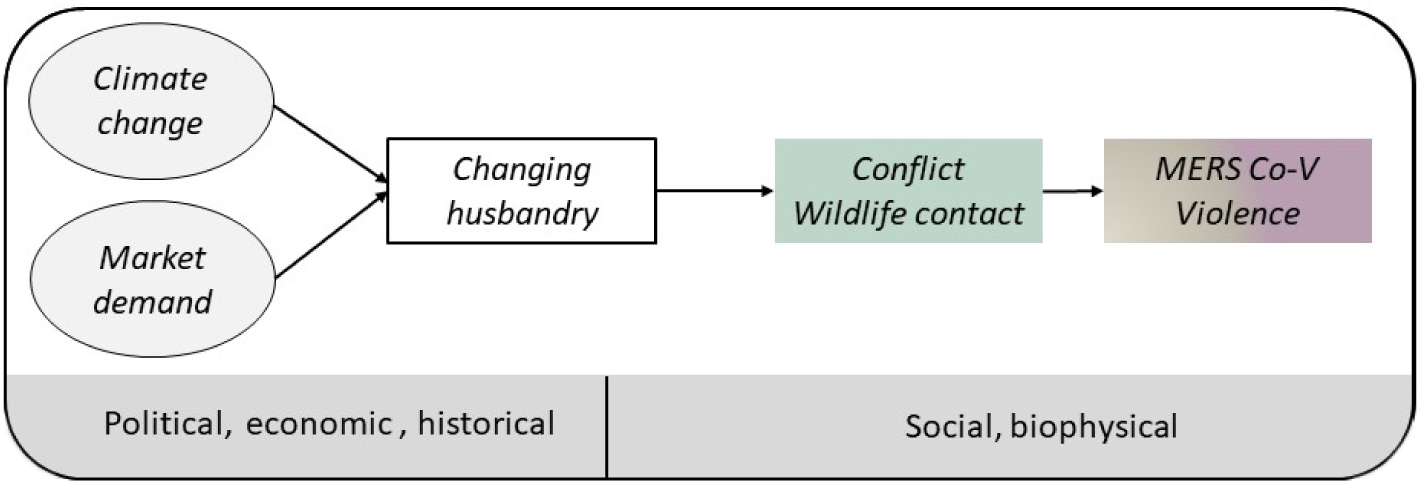
Conceptual model developed for Case Study 3. Among pastoralist communities in Ethiopia, increasing market demand and climate change, which are shaped by political and economic contexts, are changing camel husbandry. Camel husbandry has deep historical roots, and shifts in husbandry intersect with social and biophysical contexts to drive conflict and increased wildlife contact, which ultimately impact both human and animal health.

*Case study 3: MERS-CoV in Ethiopia* Case Study 3 examines the impact of human-domesticated animal, domesticated animal-wildlife, and human-wildlife relationships and interaction on zoonotic disease emergence and transmission in the larger social, cultural, economic, political and environmental contexts. Historically, semi-pastoralists of specific ethnicities dominated the camel markets and most camel trade was for local and national consumption. The demand for camel products by wealthier countries increased exponentially following MERS-CoV outbreaks in the Arabian peninsula. In response semi-pastoralists have faced increased demand for their services, and are competing with other ethnicities perceived to have social, and economic advantages. Concurrent with these shifting market forces, changes in precipitation and temperature due to climate change are leading herders to graze camels in protected areas, bringing them into contact with wildlife and increasing the risk of disease transmission—MERS-CoV and others—between humans, camels and wildlife. Finally, camel husbandry has become dangerous in some areas due to political and cultural unrest.

## DISCUSSION

We present here a systematic review of One Health conceptual models and theoretical frameworks which seek to re-conceptualize One Health in a more-than-biomedical light. We then place the findings of this review in the context of Relational One Health, and review its theoretical foundations. Finally, we present three case studies which operationalize the Relational One Health theoretical framework for research.

Recent decades have seen an increase in critically-engaged One Health frameworks which reject a reductionist view of health as a solely biomedical phenomenon. The Relational One Health theoretical framework is in many ways complementary with these earlier frameworks, being inspired by One Health of Peripheries (10) and Structural One Health (15), and being well-suited to expanding the application of the syndemic approach proposed by Rock et al. (13), and to application of the methods proposed by Woldehanna et al. (8). However, none of these earlier frameworks have succeeded in gaining broad traction within the One Health community. There are several likely explanations for this that originate in the dispersed nature of One Health, and its disciplinary foundations within fields that are proudly “purely scientific,” and may not be open to critical theory. However, there are also explanations that originate in the frameworks themselves: namely, these frameworks were either not intended for research, or only a select type of research question. We present the Relational One Health theoretical framework as a valuable addition, which can guide all One Health inquiries interested in knowledge generation regarding the determinants of multispecies health. As demonstrated through our case studies, this framework can be used to develop conceptual models which can guide and be validated through research.

Importantly, Relational One Health does not suggest that research which is solely biomedical in scope is not One Health. In such cases, Relational One Health can still be used to triangulate findings across research studies and identify gaps for further research. Relational One Health also does not prescribe a certain definition of health, which could encompass the absence of a given disease, the presence of social wellbeing or political agency, the maintenance of biodiversity, etc. The implication of this is that Relational One Health can serve as the theoretical foundation for all One Health inquiry, increasing the impact of One Health research by orienting the findings of individual studies within a shared theoretical framework. Relatedly, despite its many problems, we choose to retain the term “environment” in this framework—where it refers to the contexts in which humans, animals, and ecosystems reside and embody—to align with existing definitions of One Health, signaling that this framework is intended to be an addition to the discipline, rather than a departure from it. This framework can also serve as the basis for a more holistic examination of the intersection between One Health and critical social theory such as feminist care theory, posthumanism, and anti-colonial theory, the subject of a recent review by Van Patter et al. (19).

The wicked problems to which One Health is most suited—pandemics of zoonotic origin, neglected zoonotic diseases, climate change, and biodiversity collapse—cannot be addressed through solely biomedical counter measures or ministry of health-level programs; instead, they require large-scale political and social change. Empirical research can play a role in creating social change, but research holds greater value and impact when it can speak holistically, rather than in independent parts, and a shared theoretical foundation is critical to achieving this end. This approach has far reaching consequences, from the way the research questions are framed, to who is included in the research team, and which research traditions and languages are integrated and how. With climate change and pandemics looming, the scientific community will have to adapt its scientific tools and theories not only to the rapid pace of events, but also to the interrelatedness of phenomena surrounding us, and Relational One Health positions One Health to lead this charge.

## Data Availability

This article presents a scoping review, thus all relevant data are included in the manuscript itself

## REFERENCES

1. One Health High-Level Expert Panel (OHHLEP), Adisasmito WB, Almuhairi S, Behravesh CB, Bilivogui P, Bukachi SA, et al. One Health: A new definition for a sustainable and healthy future. Dvorin JD, editor. PLOS Pathog. 2022 Jun 23;18(6):e1010537.

2. Amuasi JH, Lucas T, Horton R, Winkler AS. Reconnecting for our future: The Lancet One Health Commission. The Lancet. 2020 May;395(10235):1469–71.

3. Horton R, Beaglehole R, Bonita R, Raeburn J, McKee M, Wall S. From public to planetary health: a manifesto. The Lancet. 2014 Mar;383(9920):847.

4. Lerner H, Berg C. A Comparison of Three Holistic Approaches to Health: One Health, EcoHealth, and Planetary Health. Front Vet Sci. 2017;4:163.

5. Davis MF, Rankin SC, Schurer JM, Cole S, Conti L, Rabinowitz P, et al. Checklist for One Health Epidemiological Reporting of Evidence (COHERE). One Health. 2017 Dec;4:14–21.

6. Page MJ, McKenzie JE, Bossuyt PM, Boutron I, Hoffmann TC, Mulrow CD, et al. The PRISMA 2020 statement: an updated guideline for reporting systematic reviews. BMJ. 2021 Mar 29;n71.

7. ResearchRabbit [Internet]. [cited 2023 Sep 14]. ResearchRabbit. Available from: https://www.researchrabbit.ai

8. Woldehanna S, Zimicki S. An expanded One Health model: integrating social science and One Health to inform study of the human-animal interface. Soc Sci Med 1982. 2015 Mar;129:87–95.

9. Zinsstag J, Schelling E, Waltner-Toews D, Tanner M. From “one medicine” to “one health” and systemic approaches to health and well-being. Prev Vet Med. 2011 Sep 1;101(3):148– 56.

10. Baquero OS. One Health of Peripheries: Biopolitics, Social Determination, and Field of Praxis. Front Public Health [Internet]. 2021 [cited 2023 Jun 21];9. Available from: https://www.frontiersin.org/articles/10.3389/fpubh.2021.617003

11. Hardy E, Standley CJ. Identifying intersectional feminist principles in the One Health framework. One Health Amst Neth. 2022 Dec;15:100404.

12. Lerner H, Berg C. The concept of health in One Health and some practical implications for research and education: what is One Health? Infect Ecol Epidemiol. 2015 Jan 1;5(1):25300.

13. Rock M, Buntain BJ, Hatfield JM, Hallgrímsson B. Animal–human connections, “one health,” and the syndemic approach to prevention. Soc Sci Med. 2009 Mar;68(6):991–5.

14. Sleeman JM, Richgels KLD, White CL, Stephen C. Integration of wildlife and environmental health into a One Health approach. Rev Sci Tech Int Off Epizoot. 2019 May;38(1):91–102.

15. Wallace RG, Bergmann L, Kock R, Gilbert M, Hogerwerf L, Wallace R, et al. The dawn of Structural One Health: a new science tracking disease emergence along circuits of capital. Soc Sci Med. 2015;

16. Hermesh B, Rosenthal A, Davidovitch N. Rethinking “One Health” through Brucellosis: ethics, boundaries and politics. Monash Bioeth Rev. 2019 Oct;37(1–2):22–37.

17. Hermesh B, Rosenthal A, Davidovitch N. The cycle of distrust in health policy and behavior: Lessons learned from the Negev Bedouin. Clegg S, editor. PLOS ONE. 2020 Aug 20;15(8):e0237734.

18. Krieger N. Ecosocial Theory of Disease Distribution: Embodying Societal & Ecologic Context. In: Epidemiology and the People’s Health. New York, NY: Oxford University Presss; 2011. p. 202–35.

19. Van Patter LE, Linares-Roake J, Breen AV. What does One Health want? Feminist, posthuman, and anti-colonial possibilities. One Health Outlook. 2023 Mar 10;5(1):4.

